# Interferon pathway lupus risk alleles modulate risk of death from acute COVID-19

**DOI:** 10.1101/2021.11.01.21265766

**Authors:** Ilona Nln, Ruth Fernandez-Ruiz, Theresa L. Wampler Muskardin, Jacqueline L. Paredes, Ashira D. Blazer, Stephanie Tuminello, Mukundan Attur, Eduardo Iturrate, Christopher M. Petrilli, Steven B. Abramson, Aravinda Chakravarti, Timothy B. Niewold

**Affiliations:** Colton Center for Autoimmunity, NYU Grossman School of Medicine, New York, NY; Center for Human Genetics and Genomics, NYU Grossman School of Medicine, New York, NY; Divison of Rheumatology, Department of Medicine, NYU Grossman School of Medicine, New York, NY; Department of Medicine, NYU Grossman School of Medicine, New York, NY

## Abstract

Type I interferon (IFN) is critical in our defense against viral infections. Increased type I IFN pathway activation is a genetic risk factor for systemic lupus erythematosus (SLE), and a number of common risk alleles contribute to the high IFN trait. We hypothesized that these common gain-of-function IFN pathway alleles may be associated with protection from mortality in acute COVID-19. We studied patients admitted with acute COVID-19 (756 European-American and 398 African-American ancestry). Ancestral backgrounds were analyzed separately, and mortality after acute COVID-19 was the primary outcome. In European-American ancestry, we found that a haplotype of interferon regulatory factor 5 (IRF5) and alleles of protein kinase cGMP-dependent 1 (PRKG1) were associated with mortality from COVID-19. Interestingly, these were much stronger risk factors in younger patients (OR=29.2 for PRKG1 in ages 45-54). Variants in the IRF7 and IRF8 genes were associated with mortality from COVID-19 in African-American subjects, and these genetic effects were more pronounced in older subjects. Combining genetic information with blood biomarker data such as C-reactive protein, troponin, and D-dimer resulted in significantly improved predictive capacity, and in both ancestral backgrounds the risk genotypes were most relevant in those with positive biomarkers (OR for death between 14 and 111 in high risk genetic/biomarker groups). This study confirms the critical role of the IFN pathway in defense against COVID-19 and viral infections, and supports the idea that some common SLE risk alleles exert protective effects in anti-viral immunity.

**Background:** We find that a number of IFN pathway lupus risk alleles significantly impact mortality following COVID-19 infection. These data support the idea that type I IFN pathway risk alleles for autoimmune disease may persist in high frequency in modern human populations due to a benefit in our defense against viral infections.

**Translational Significance:** We develop multivariate prediction models which combine genetics and known biomarkers of severity to result in greatly improved prediction of mortality in acute COVID-19. The specific associated alleles provide some clues about key points in our defense against COVID-19.

## Introduction

The severity of COVID-19 infection is strikingly variable between individuals, ranging from asymptomatic infection to cytokine storm with respiratory insufficiency and multiorgan failure and death. While some risk factors have been identified, it is clear that these factors cannot fully explain the variation in severity between individuals. Type I interferon (IFN) is a critical viral defense pathway, and studies have documented that rare knockout loss-of-function genetic variants and autoantibodies against type I IFNs are associated with severe COVID-19 (*1, 2)*. We have studied the genetics of the type I IFN system in the autoimmune disease systemic lupus erythematosus (SLE) (*3-6*). Via this work, we have found a number of common genetic polymorphisms that result in increased type I IFN production and signaling in humans (*3, 6*). These gain-of-function polymorphisms are risk factors for SLE and other autoimmune diseases (*7*). We hypothesized that these SLE-associated polymorphisms that result in increased type I IFN pathway activation would reduce the severity and mortality of acute COVID-19.

Many of the risk alleles associated with autoimmune disease are common ancestral alleles, which raises the question of whether there may be some benefit to the individual to explain their persistence at high frequency in the population. One possibility is that these gain-of-function alleles in the IFN pathway offer some protection from viral infections. In this scenario, the common polymorphisms may offer some overall protection against viral infection that is beneficial to the population, but the uncommon situation in which someone carries a large number of these alleles or an unlucky synergistic combination would result in risk of autoimmunity. This rare case of autoimmunity may be outweighed by a more common benefit to the immune system with respect to viral infection. While this theory is attractive, it has been hard to test definitively in human populations, because common viral exposures in the population can be difficult to track and quantify. The severe COVID-19 outbreak in New York City in 2020 provided us with a unique opportunity to study how genetic variation in the IFN pathway impacts the outcome of acute viral illness. In this study, we find general support for the hypothesis that genetic variations that are protective against SLE or associated with reduced type I IFN responses are associated with an increased mortality in acute COVID-19. Additionally, we develop multivariate prediction models which combine genetics and known biomarkers of severity to result in greatly improved prediction of mortality in acute COVID-19.

## Methods

### Subjects

We collected genomic DNA from 1149 COVID-19 positive patients: European-American n=756 (n=144 deceased, n=612 alive), and African-American n=398 (n=63 deceased, n=335 alive). The mean (standard deviation) age and body mass index (BMI) for the cohort were 67.72 (17.86) years and 34.96 (15.7) respectively. 46.9% of the subjects were female. All subjects tested positive for COVID-19 by PCR test, and samples were obtained at the time of clinical care. Mortality was defined as in-hospital mortality during that admission for COVID-19. C-reactive protein (CRP), troponin, and D-dimer were measured in the NYU Clinical Laboratory and were abstracted from the medical record. All subjects provided informed consent and the study was approved by the NYU Institutional Review Board.

### Genotyping

We studied SNPs in the following IFN pathway genes in which gain-of-function properties in humans have been previously documented (*4, 5, 8-14*): IRF5 - rs2004640, rs3807306, rs10488631, rs2280714; IRF7 -rs702966, rs4963128, rs1131665; IRF8 - rs17445836, rs12444486; STAT4 - rs7574865; PRKG1 rs7897633, and IFIH1 - rs1990760. We used a low coverage whole genome sequencing strategy in this study (*15*). After quality control, low coverage data were used to impute all common (minor allele frequency >1%) human polymorphism genotypes for each sample, using reference populations from the 1000 Genomes data sets. We selected the above SNPs from this larger dataset for analysis in this study.

### Statistical analysis

Our primary analysis was to compare mortality from COVID-19 between genotype groups for the various IFN pathway SLE risk alleles. This was done using logistic regression models using age, sex, and BMI as covariates. When a significant interaction with a covariate was detected, this was explored and modeled further to understand whether a particular subgroup was more or less affected than the group as a whole. For example, significant interaction with age resulted in age-group analyses to detect the associated patterns in each ancestral background. No statistical interaction was found for BMI or sex in either ancestry. Each analysis was performed in each ancestral background separately first, and there were no shared signals that allowed for meta-analysis. We used a threshold p-value of 0.05 to screen for SNPs to take into further modeling as described below.

In secondary analyses, we examined blood biomarkers of severe COVID-19, including D-dimer, troponin, and CRP levels in the context of the same genotypes to generate overall predictive models (*16-18*). Given the significant multi-factor interdependencies, we utilized regression models to detect interactions and reduce redundancy due to correlated variables. Although ideally a more granular age group breakdown could be informative, due to our limited sample size, we focus on two major age groups (<55 years old and >=55 years old), which we found is sufficient to highlight patterns that apply to younger vs. older cohorts. For the biomarkers, we classify each subject into a low or high group depending on whether the biomarker level is below or above an ancestry-specific threshold respectively. The ancestry-specific threshold is computed as the average of the two medians of the deceased and non-deceased distributions for each ancestry, as different ancestral backgrounds had different biomarker distributions. These data were taken into combinatorial factor analyses along with the associated risk genotypes identified in each ancestral background above.

## Results

### European-American ancestry genetic associations with COVID-19 mortality

IRF5 haplotypes are associated with graded SLE risk, with examples of protective, neutral, and risk haplotypes that span the gene and downstream region (*12*). Given this, we modeled IRF5 genotypes as haplotype combinations. We observed that carriage of a specific IRF5 haplotype which is associated with lower serum IFN in SLE patients was strongly predictive of mortality related to COVID19 in patients of European-American ancestry (OR=2.25, p=0.014, Figure 1A). This allelic association was specific to European ancestry, similar to previous IRF5 genetic association studies in autoimmune disease, which have documented significant differences in associations between ancestral backgrounds (*19*). Interestingly, this genetic effect was strongest in younger patients, particularly in the 45-54 year old group (Figure 1A).

**Figure 1.**
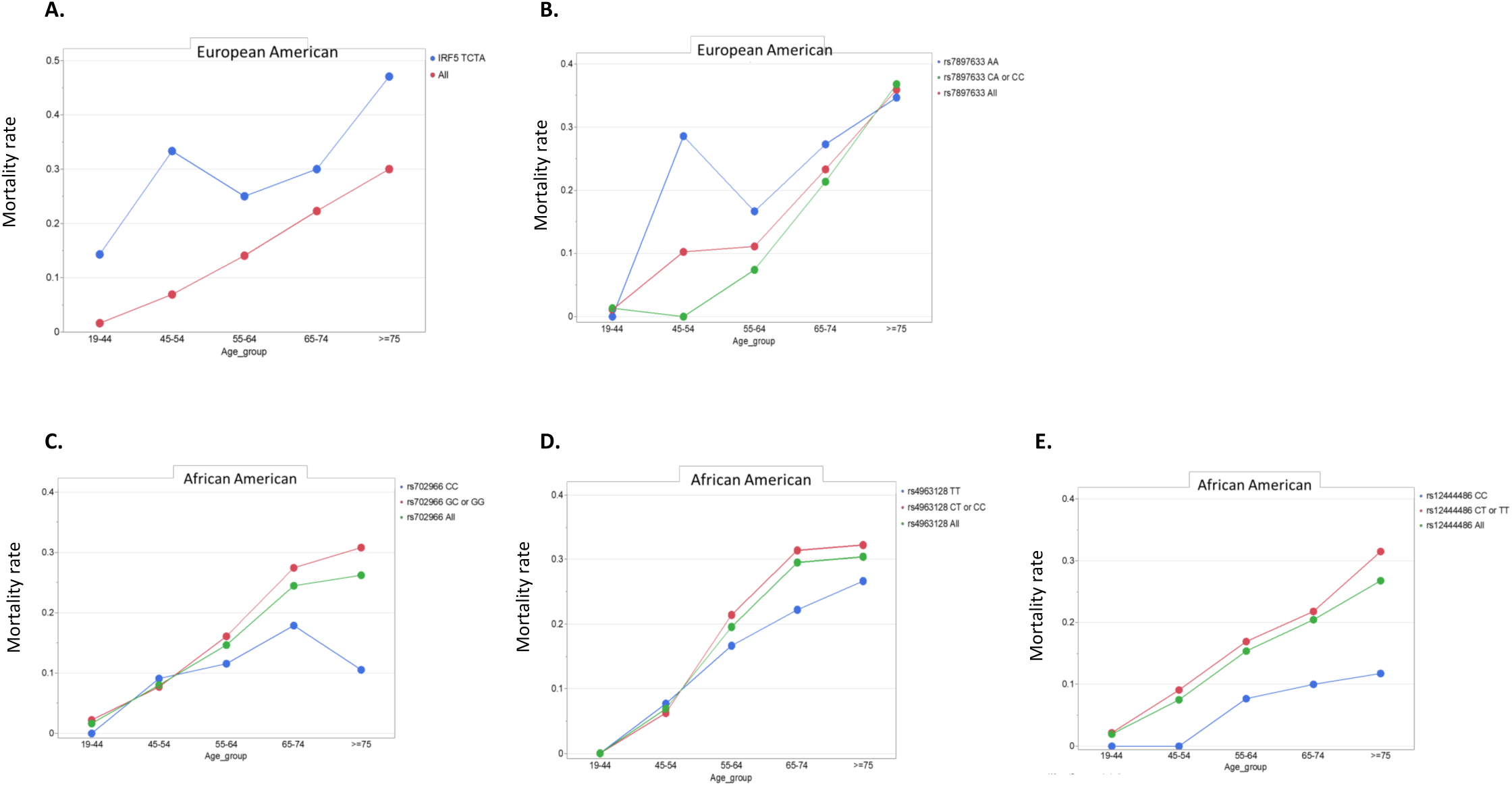
COVID-19 mortality rate vs. age stratified by genetic variants. A. shows data for the IRF5 TCTA haplotype in European-Americans, in which TCTA indicates the alleles at the rs2004640, rs3807306, rs10488631, and rs2280714 SNPs in that order, B. shows PRKG1 rs7897633 genotype in European-Americans, C. shows rs702966 genotype in African-Americans, D. shows rs4963128 genotype in African-Americans, and E. shows rs12444486 genotype in African-Americans. Mortality rate shown ranges from 0 to 1, with 0.2 corresponding to 20% mortality for example.

Alleles of PRKG1 were also associated with mortality from COVID-19 in the European-American ancestry cohort (OR=1.80, p=0.0057), and this risk factor was very strong in younger patients (OR=29.2, p=0.01 in ages 45-54 for homozygous AA genotype) (Figure 1B). In this group, mortality was over 25%, as compared to less than 5% in all subjects ages 45-54. We did not observe associations between this SNP and COVID-19 mortality in African-American subjects. While this could be a power issue, in the previous study of this SNP in lupus patients the association with type I IFN was only observed in European-American lupus patients, and not in African-American patients in whom the prevalence of the SNP was slightly higher (*3*). This could suggest a causal element in the region that is tagged by this SNP allele in European but not African ancestry chromosomes.

### African-American ancestry genetic associations with COVID-19 mortality

In African-American subjects, we found that the G allele of the IRF7 SNP rs702966 was associated with mortality in COVID-19 patients (OR=2.09, p=0.015, Figure 1C). We have previously shown that this allele is associated with decreased circulating type I IFN in African-American SLE patients (*8*). We also found that the rs4963128 SNP in the adjacent PHRF1 gene that has been associated with SLE and type I IFN levels in SLE (*8*) was associated with differences in mortality in acute COVID-19. The TT genotype of rs4963128 was associated with protection from COVID-19 mortality (OR=0.52 p=0.014, Figure 1D), and this genotype is associated with higher type I IFN levels in African-American SLE patients (*8*). It seems likely that the PHRF1 SNP could tag a functional genetic element that modulates the adjacent IRF7 gene to influence type I IFN, as PHRF1 is not thought to function in the type I IFN pathway. When we combined data from these two genotypes at the IRF7 locus and designate carriers of one or more risk allele at either of the two SNP positions as risk genotype, the OR for mortality during acute COVID-19 increased to 2.88 (p=0.008). Both of these IRF7 alleles demonstrate a greater effect upon mortality in older subjects as compared to younger subjects (Figure 1C and D).

IRF8 genotype at rs12444486 was associated with protection from mortality in COVID-19 in African-American subjects (OR=0.38, p=0.047, Figure 1E). This association was not observed in European ancestry subjects. Interestingly, another lupus-associated IRF8 allele was tested (rs17445836), but this SNP was not associated with COVID-19 mortality in either ancestral background. Previous studies in autoimmune diseases have supported multiple independent signals in the IRF8 locus with evidence for different functional associations (*4, 20*).

Some interesting patterns emerge in these two populations. First, the data fit an overall model in which SNPs associated with higher IFN in SLE patients are associated with lower mortality in acute COVID-19. This is illustrated in the correlation plot in Figure 2, demonstrating the correlation between type I IFN ratio between genotype groups in SLE patients as compared to the OR for COVID-19 mortality between genotype groups (r = 0.937, p=0.019). This trend is consistent across the various genetic polymorphisms and between ancestral backgrounds. Comparing the findings in European-American vs. African-American subjects, there are differences in the age groups that are most impacted by genotype, as well as the specific polymorphisms associated, as shown in Figure 1. This suggests diversity in regulation of the IFN pathway between ancestral backgrounds, both with respect to the particular genetic elements as well as age-related differences.

**Figure 2.**
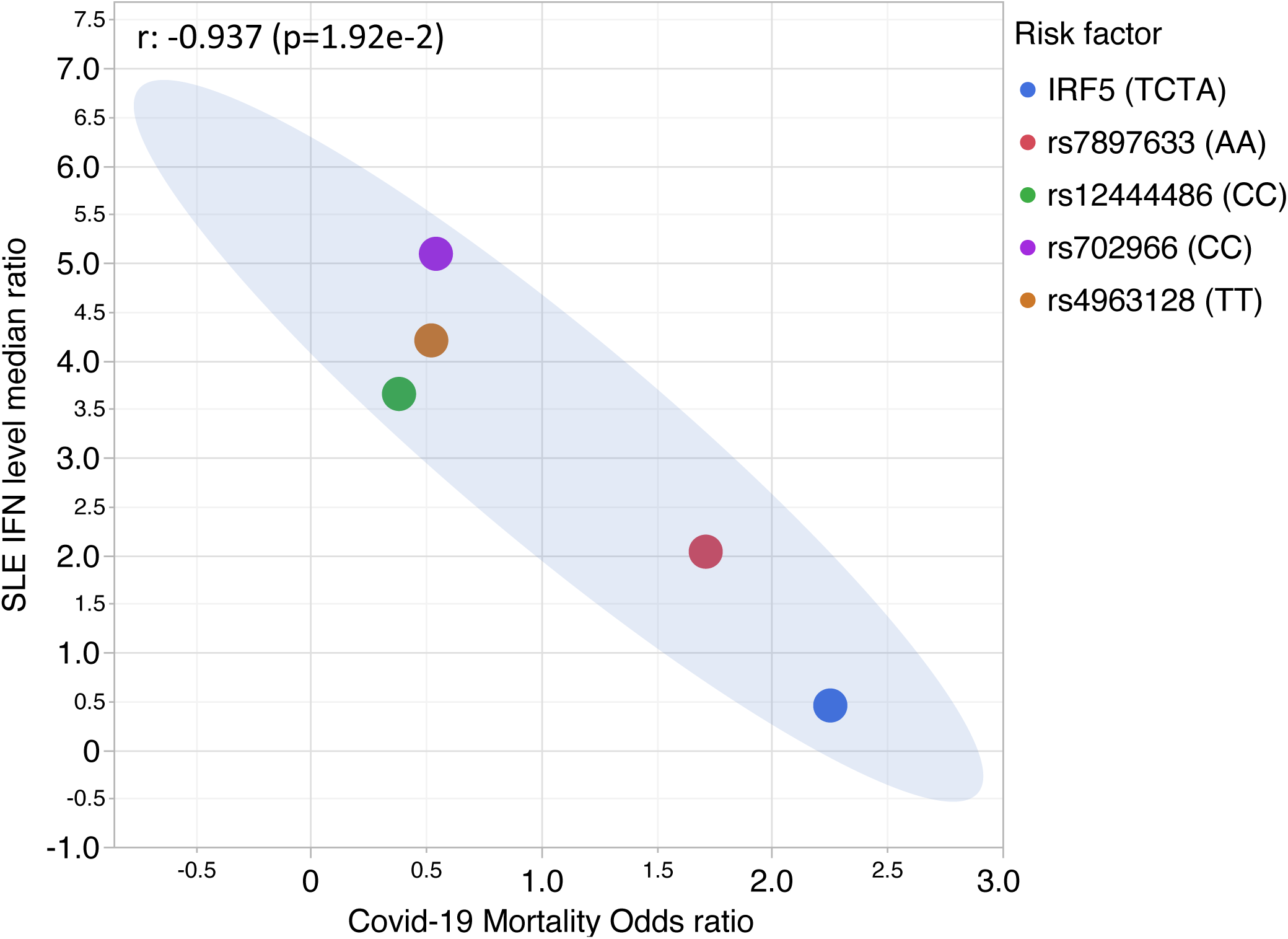
Correlation plot between type I IFN ratio by genotype and odds ratio for mortality related to acute COVID-19 by the same genotype. The IFN ratios are calculated from our previous published studies in SLE noted in the Methods section, computing a ratio of median circulating IFN values between the genotype groups. Genotype categories used are the same for both the IFN analysis and the COVID-19 mortality analysis. 95% confidence is shown with the blue shading. TCTA indicates carriage of the associated IRF5 COVID-19 risk haplotype.

### Gene and biomarker risk factor interactions – European-American ancestry results

We next analyzed the IFN pathway genotype data in the context of biomarkers that are known to be associated with mortality in acute COVID-19, including age, D-Dimer, CRP, and troponin (*16-18*). We designated risk genotype as either PRKG1 rs7897633 AA homozygous or IRF5 TCTA haplotype carrier (Figure 3). Strikingly, we did not see any deaths in European-American subjects under age 55 who did not have one of the two risk gene polymorphisms (IRF5 or PRKG1). In contrast, presence of one of the risk genotypes was associated with a large OR for mortality in the absence of biomarkers for severity (OR>10), and this increased greatly as the number of biomarkers increased. In older subjects, the impact of genotype was smaller and the overall chance of mortality due to acute COVID-19 increased. In subjects over age 55 lacking one of the two risk genotypes, there was a gradual increase in mortality with a greater number of positive biomarkers, fitting a linear model. In those with one or both of the risk genotypes, the OR for mortality was higher than those lacking risk genotypes, and the OR increased with increasing biomarker positivity in a log2 growth pattern. Subjects that carry risk genes and risk biomarkers have a significant increase in risk of COVID-19 mortality (>95% confidence following Bonferroni correction for family-wise type I error).

**Figure 3.**
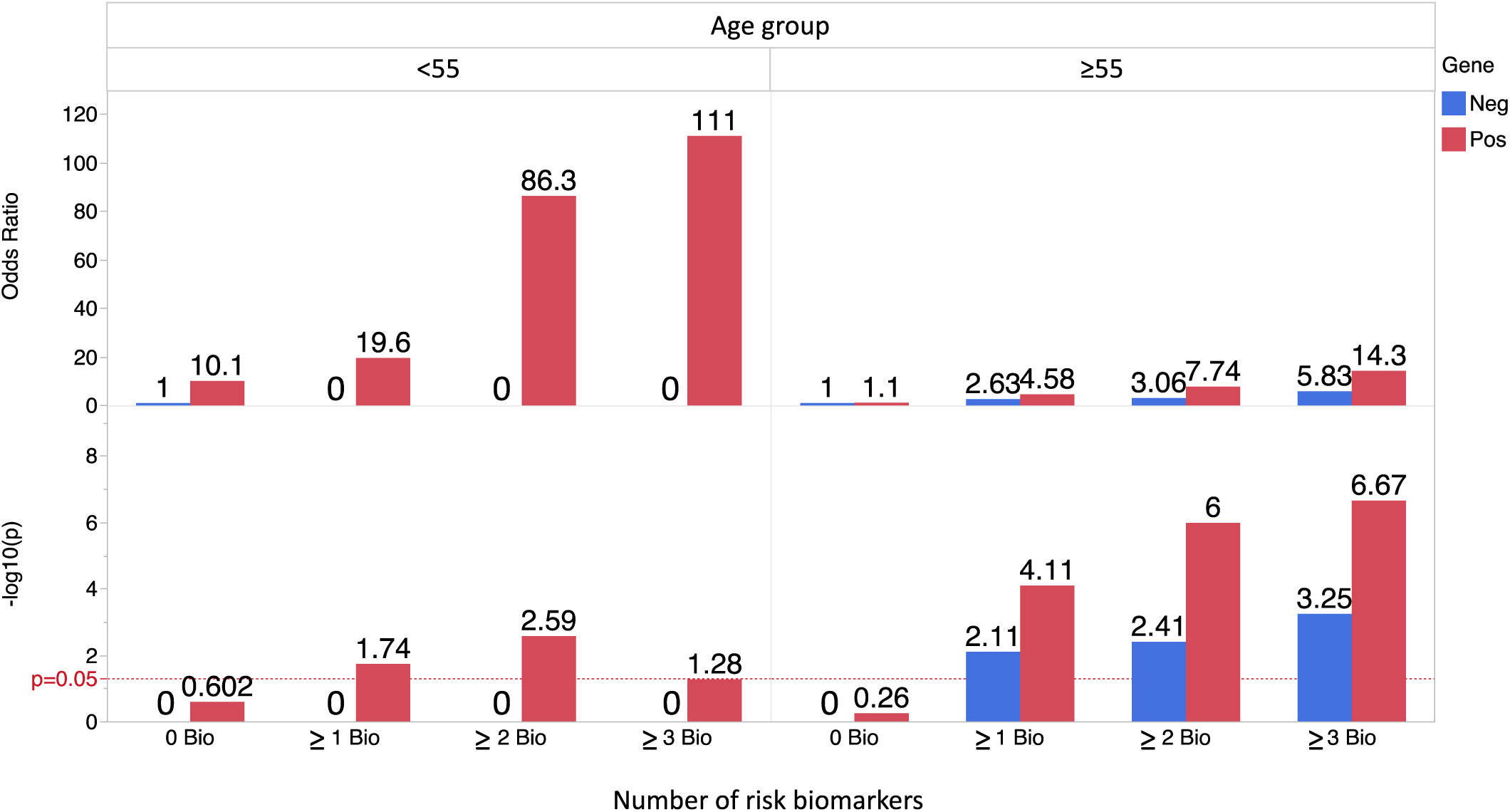
Association analysis results for European ancestry cohort, showing odds ratios for mortality from acute COVID-19 in older (>=55 years) and younger (<55 years) subjects and in the presence or absence of biomarkers for severe disease. The genetic risk factors (coded as “Pos” and shown as red bars) are rs7897633 AA homozygosity and TCTA IRF5 haplotype carriers, subjects lacking these two genotypes are coded as “Neg” and shown as blue bars. The biomarker risk factors include D-Dimer>510 ng/mL, CRP>110 mg/L, and Troponin>0.015 ng/mL, and each biomarker above these thresholds was weighted as 1 positive biomarker. All odds ratios and hypothesis test p-values were calculated for mortality risk with respect to the lowest risk group (gene negative and biomarker negative). Dashed lines showing the minimum threshold for 95% statistical confidence are shown in red for each age group (row). When there are no deceased subjects in a given category, no OR and p-value are shown.

Table 1 shows the breakdown of mortality risk associated with individual biomarkers and all of the possible combinations, with and without genetic risk factors. When examined separately or in conditional models, each of the 3 biomarker risk factors exerted effects which were independent from each other (Figure 4). As expected, risk was highest when all 3 biomarkers were present.

**Table 1.** Association analysis results for European ancestry cohort stratified by age and number of biomarkers. The gene risk factors are rs7897633 AA homozygous allele and TCTA IRF5 haplotype carriers. The biomarker risk factors include D-Dimer>510 ng/mL, CRP>110 mg/L, and Troponin>0.015 ng/mL. All odds ratios and hypothesis test p-values were calculated for mortality risk with respect to the lowest risk group (Gene 0 and biomarker negative), reflected in the OR of 1 for these groups in the table. – means that no deceased subjects were present in that category.

| Age (yrs) | Risk factor | OR (95% CI) | p-value | Deceased |  |  | Alive |  |  |
| --- | --- | --- | --- | --- | --- | --- | --- | --- | --- |
|  |  |  |  | N Risk factor + | N Risk factor - | Freq of risk factor+ | N Risk factor + | N Risk factor - | Freq of risk factor+ |
| <55 | Genes & 3 Bio | 111.00 (1.57-7848.40) | $5.3 \times 10^{-2}$ | 1 | 4 | 0.20 | 0 | 94 | 0.00 |
| | Genes & $\geq 2$ Bio | 86.33 (2.89-2582.92) | $2.6 \times 10^{-3}$ | 3 | 2 | 0.60 | 1 | 93 | 0.01 |
| | Genes & $\geq 1$ Bio | 19.59 (0.94-406.47) | $1.8 \times 10^{-2}$ | 4 | 1 | 0.80 | 8 | 86 | 0.09 |
| | Genes & 0 Bio | 10.09 (0.36-284.53) | $2.5 \times 10^{-1}$ | 1 | 4 | 0.20 | 5 | 89 | 0.05 |
|  | 0 Gene & 0 Bio | 1 | 1 | 0 | 5 | 0.00 | 18 | 76 | 0.19 |
| | 0 Gene & $\geq 1$ Bio | - | - | 0 | 5 | 0.00 | 46 | 48 | 0.49 |
| | 0 Gene & $\geq 2$ Bio | - | - | 0 | 5 | 0.00 | 17 | 77 | 0.18 |
|  | 0 Gene & 3 Bio | - | - | 0 | 5 | 0.00 | 3 | 91 | 0.03 |
| $\geq 55$ | Genes & 3 Bio | 14.25 (4.92-41.27) | $2.1 \times 10^{-7}$ | 19 | 117 | 0.14 | 10 | 416 | 0.02 |
| | Genes & $\geq 2$ Bio | 7.74 (3.19-18.81) | $1.0 \times 10^{-6}$ | 32 | 104 | 0.24 | 31 | 395 | 0.07 |
| | Genes & $\geq 1$ Bio | 4.58 (2.00-10.49) | $7.8 \times 10^{-5}$ | 44 | 92 | 0.32 | 72 | 354 | 0.17 |
| | Genes & 0 Bio | 1.10 (0.33-3.64) | $5.4 \times 10^{-1}$ | 5 | 131 | 0.04 | 34 | 392 | 0.08 |
|  | 0 Gene & 0 Bio | 1 | 1 | 8 | 128 | 0.06 | 60 | 366 | 0.14 |
| | 0 Gene & $\geq 1$ Bio | 2.63 (1.20-5.76) | $7.7 \times 10^{-3}$ | 70 | 66 | 0.51 | 200 | 226 | 0.47 |
| | 0 Gene & $\geq 2$ Bio | 3.06 (1.34-6.98) | $3.8 \times 10^{-3}$ | 40 | 96 | 0.29 | 98 | 328 | 0.23 |
| | 0 Gene & 3 Bio | 5.83 (2.11-16.11) | $5.6 \times 10^{-4}$ | 14 | 122 | 0.10 | 18 | 408 | 0.04 |

**Figure 4.**
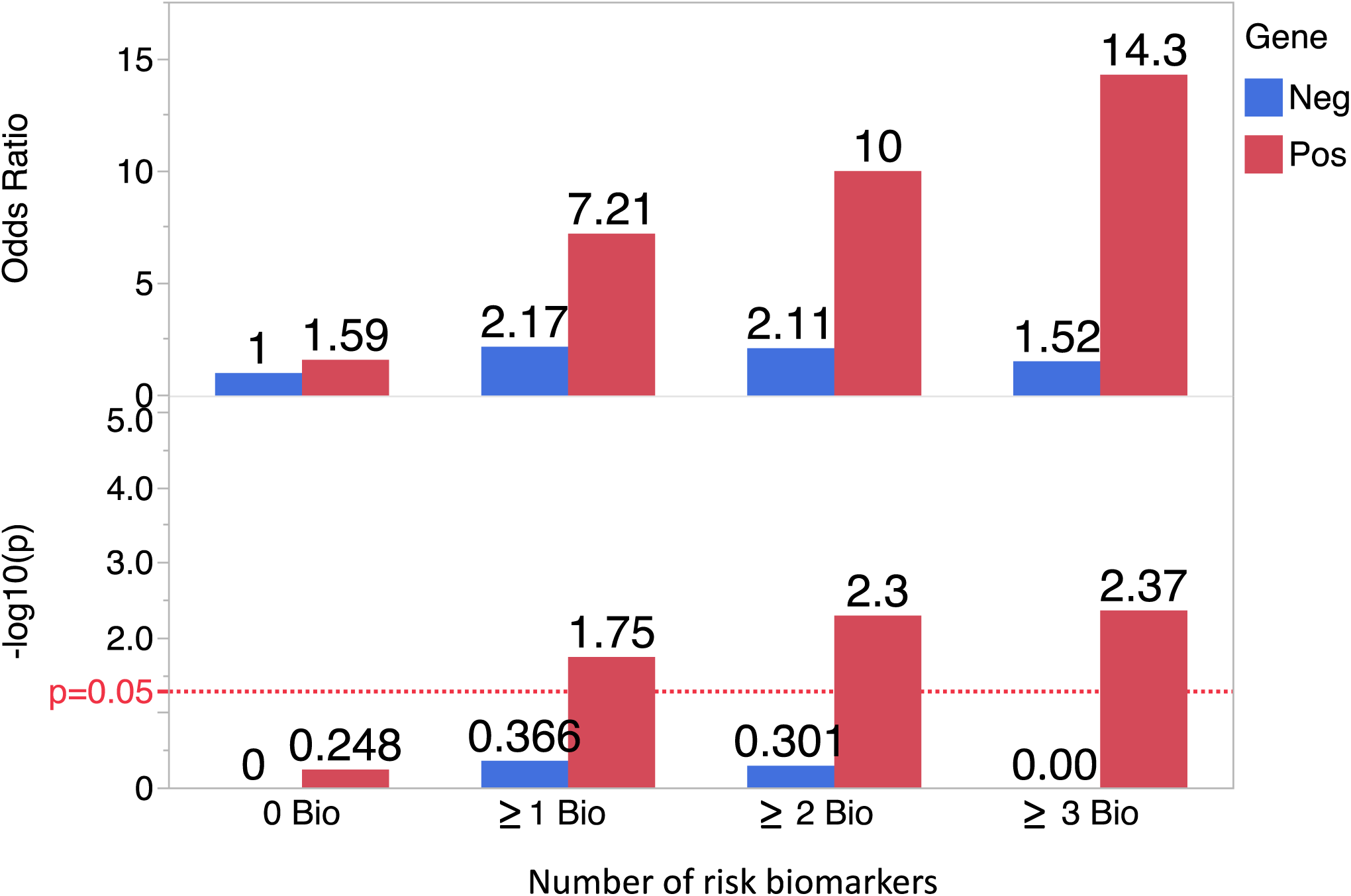
Association analysis results for African-American ancestry cohort, showing odds ratios for mortality from acute COVID-19 in the presence or absence of biomarkers for severe disease. The genetic risk factors (coded as “Pos” and shown as red bars) are IRF7 region rs4963128 C allele carriers and rs702966 G allele carriers. Subjects who do not carry any one of these alleles are coded as “Neg” and shown as blue bars. The biomarker risk factors include D-Dimer>560 ng/mL, CRP>115 mg/L, and Troponin>0.01 ng/mL, and each biomarker above these thresholds was weighted as 1 positive biomarker. All odds ratios and hypothesis test p-values were calculated for mortality risk with respect to the lowest risk group (gene negative and biomarker negative). Dashed lines showing the minimum threshold for 95% statistical confidence are shown in red.

### Gene and biomarker risk factor interactions – African-American ancestry results

In the African-American ancestry cohort, we considered carriage of either of two COVID risk SNPs in the IRF7 region (rs702966 G or rs4963128 T) as a risk genotype carrier. IRF8 genotype was initially included in the model as well, although it did not confer additional predictive capacity and dropped out, with the two IRF7 SNPs providing the best model fit. Similar to the European-American analysis above, we found significant risk of mortality due to acute COVID-19 in those with risk genotypes and positive biomarkers. In the African-American cohort, we did not see a statistically significant increase in risk in those with risk genotypes but no positive biomarkers (Figure 4). Because the group was smaller, we were not able to see significant patterns when subjects were split into <55 years old vs. ≥ 55 years old.

## Discussion

We document multiple associations between lupus-associated functional polymorphisms in the type I IFN pathway and mortality from acute COVID-19. This study supports the concept that type I IFN is critical in our defense against COVID-19, and that both rare loss-of-function alleles (*1*) and common gain-of-function alleles tune the type I IFN pathway in ways that impact our viral defense. This model is also supported by recent studies which show that genetic variants and increased circulating levels of the IFN-induced 2’-5’-oligoadenylate synthetase 1 (OAS1) modulate risk of severe COVID-19 (*21-23*). Alleles of the OAS1 gene have also been associated with the autoimmune condition Sjogren’s disease (*24*). Sjogren’s is characterized by high IFN levels, similar to SLE. The hypothesis that common IFN pathway autoimmune disease risk alleles are present in the population at high frequency because of a beneficial effect upon viral immunity has been proposed previously, but it has been difficult to study directly due to the inherent difficulties in studying viral exposures in the human population. The COVID-19 pandemic allowed for a unique window into this phenomenon, as a previously unencountered virus swept through the human population. Whether the phenomenon we report is associated with protection from viruses in patients with lupus or autoimmune disease is not known (*25*), but this could be confounded by the immunosuppressive treatments often used in these conditions.

Interestingly, some but not all of the type I IFN functional alleles were associated with differential outcomes following COVID-19. While this could relate to statistical power issues to some degree, the stark differences between SNP associations across ancestral backgrounds likely indicates diversity in associations. This study provides some insight into which parts of the type I IFN pathway are important in defense against COVID-19, and in which populations. Also, these data could indicate which parts of the IFN pathway are associated with lupus but not COVID-19 mortality. This may suggest which parts of the exuberant type I IFN response in lupus may be more safely targeted, at least with respect to COVID-19.

Combining our genetic results with known biomarkers for COVID-19 mortality greatly improved our predictive models, including the finding that there was no mortality in younger European-American subjects who lacked the risk genotypes. While this would benefit from independent replication, our results are independently significant and striking, and could be of clinical utility in the younger patient population. The alleles we report have not been discovered in large COVID-19 genetic screens conducted thus far (*22, 23, 26-28*), but that is likely related to a number of factors. Our focused candidate gene study requires less type I error correction than a whole genome screen, and allows for detailed investigation of interactions with age and biomarkers, which was critical in refining our association models. Our single center study has less variation and heterogeneity with respect to case identification, treatment protocols, and demographic and environmental factors than large multi-center studies. Also, our study includes African-American subjects, while some of the other large screens to date have been limited to European ancestry, and our study supports the idea that associations will differ significantly between populations.

Our study has limitations, as the large COVID-19 wave in New York City in 2020 occurred, many subjects were recruited but we have not been able to assemble a large replication cohort as case numbers in New York City have remained low after the severe first wave. A replication cohort would be ideal, and we will await with interest future studies of these loci from data sets at other centers. This is a candidate gene study, which allows us to benefit from prior knowledge and rationale, and we maintain an appropriate type I error correction. Despite this, replication in future studies will be important.

In summary, we document a number of IFN pathway lupus risk alleles that significantly impact mortality following COVID-19 infection, and these alleles provide some clues about key points in our viral defense against COVID-19. The study also supports the idea that type I IFN pathway risk alleles for autoimmune disease may persist in high frequency in modern human populations due to a benefit in our defense against viral infections.

## Supporting information

Equator checklist

## Data Availability

All data produced in the present study are available upon reasonable request to the authors

## Abbreviations

IFN: interferon
SLE: systemic lupus erythematosus
COVID-19: coronavirus disease 2019
IRF: interferon regulatory factor,
PRKG1: protein kinase cGMP-dependent 1
CRP: C-reactive protein
STAT4: signal transducer and activator of transcription 4
IFIH1: interferon-induced with helicase c domain 1
BMI: body mass index
SNP: single nucleotide polymorphism
NYU: New York University,
OR: odds ratio

